# Ovarian carcinosarcoma is a distinct form of ovarian cancer with poorer survival compared to tubo-ovarian high grade serous carcinoma

**DOI:** 10.1101/2022.02.19.22271197

**Authors:** Robert L. Hollis, Ian Croy, Mike Churchman, Clare Bartos, Tzyvia Rye, Charlie Gourley, C. Simon Herrington

**Author notes:** Correspondence: Dr Robb Hollis, Cancer Research UK Edinburgh Centre, MRC Institute of Genetics and Cancer, University of Edinburgh, Crewe Road South, Edinburgh, EH4 2XU, Scotland, UK. **COIs:** RLH: consultancy fees from GSK outside the scope of this work. IC: none. MC: none. TR: none. CB: none. CG: grants from AstraZeneca, MSD, BMS, Clovis, Novartis, BerGenBio, Medannexin and Artios; personal fees from AstraZeneca, MSD, GSK, Tesaro, Clovis, Roche, Foundation One, Chugai, Takeda, Sierra Oncology, Takeda and Cor2Ed outside the submitted work; patents PCT/US2012/040805 issued, PCT/GB2013/053202 pending, 1409479.1 pending, 1409476.7 pending, and 1409478.3 pending. CSH: none. **Author contributions** RLH: conceptualisation, formal analysis, investigation, methodology, visualisation, funding acquisition, writing – original draft. IC: investigation, writing – review and editing. MC: data curation, project administration, writing – review and editing. TR: data curation, investigation, writing – review and editing. CB: data curation, writing – review and editing. CG: supervision, resources, writing – review and editing CSH: investigation, methodology, supervision, writing – review and editing.

## Abstract

**BACKGROUND:** Ovarian carcinosarcoma (OCS) is an uncommon, biphasic and highly aggressive ovarian cancer type, which has received relatively little research attention.

**METHODS:** We curated the largest pathologically-confirmed OCS cohort to date, performing detailed histopathological characterisation, analysis of features associated with survival, and comparison against high grade serous ovarian carcinoma (HGSOC).

**RESULTS:** 82 OCS patients were identified; overall survival was poor (median 12.7 months). 79% demonstrated epithelial components of high grade serous (HGS) type, while 21% were endometrioid. Heterologous elements were common (chondrosarcoma in 32%, rhabdomyosarcoma in 21%, liposarcoma in 2%); chondrosarcoma was more frequent in OCS with carcinomatous components of endometrioid type. Earlier stage, complete surgical resection, and treatment with platinum-containing chemotherapy were associated with prolonged survival; however, risk of relapse and mortality was high across all patient groups. Histological subclassification did not identify subgroups with distinct survival. Compared to HGSOC, OCS patients were older at diagnosis (P<0.0001), more likely to be FIGO stage I (P=0.025), demonstrated lower chemotherapy response rate (P=0.001) and had significantly poorer survival (P<0.0001).

**CONCLUSION:** OCS represents a distinct, highly lethal form of ovarian cancer for which new treatment strategies are urgently needed. Aggressive adjuvant chemotherapy should be considered for all patients, including those with early stage disease.

## BACKGROUND

Ovarian carcinosarcoma (OCS) – previously also known as mixed malignant Müllerian tumour – is an uncommon, highly aggressive cancer of the female genital tract (1). Unlike more common ovarian cancers, OCS is biphasic, comprising malignant epithelial (carcinomatous) and malignant mesenchymal (sarcomatous) populations.

While it was originally thought that OCS may represent collisions of separate carcinomas and sarcomas (2, 3), molecular studies have revealed a clonal relationship between these two malignant cell populations (4), pointing to a shared malignant ancestor cell. Much of our understanding of OCS is inferred from uterine carcinosarcoma (UCS), a more common cancer in women (5). However, it is well recognized that cancers presenting on the ovary bear stark clinical and molecular differences compared to those of the uterus that demonstrate similar histology (6-10). Indeed, limited available data suggest differences in the molecular landscape of OCS compared to UCS, though the number of comprehensively characterized OCS samples to date is extremely low (4, 11).

OCS are highly heterogeneous, defined by the presence of both high grade carcinomatous and high grade sarcomatous cell populations (12). Carcinomatous elements may be of any ovarian high grade carcinoma type (high grade serous, endometrioid, clear cell, mucinous). The sarcomatous compartment may be classified as homologous – demonstrating either non-specific appearance or differentiation native to the female genital tract – or heterologous, showing differentiation physiologically foreign to the adnexa (1, 12). The most common heterologous sarcomatous elements are chondroid (chondrosarcoma) and rhabdoid differentiation (rhabdomyosarcoma), with other heterologous elements noted only in rare cases (angiosarcoma, osteosarcoma and liposarcoma; <5% of cases) (1, 12).

Despite its aggressive behavior (13, 14), OCS has received relatively little research attention to date. A limited number of studies have characterized an appreciable number of OCS patients in detail (15-20). However, these studies have focused on describing patient outcome and typically have not performed contemporary pathology review to confirm OCS diagnosis; moreover, these studies have not described the histopathological characteristics of cases in detail.

Currently, detailed histopathological classification of the carcinomatous and sarcomatous elements is not routinely performed in OCS diagnosis. Little is therefore known about the relationship between different histopathological features of OCS, or whether these features are related to distinct clinical characteristics of OCS patients. We sought to robustly curate a large cohort of OCS cases, performing detailed clinical and histopathological characterization to improve our understanding of this highly aggressive tumour type.

## METHODS

### Cohort identification and clinical annotation

All ovarian cancer cases with a documented diagnosis of carcinosarcoma up to 31^st^ December 2020 were identified from the Edinburgh Ovarian Cancer Database (figure 1), wherein the details of diagnosis, treatment and outcome of all ovarian cancer patients treated at the Edinburgh Cancer Centre are entered prospectively as part of routine care (21). Baseline clinicopathological characteristics, treatment and outcome data were extracted. Overall survival (OS) was calculated from date of pathologically confirmed diagnosis. Progression-free survival (PFS) was calculated as time from pathologically confirmed diagnosis to recurrence/progression (supplementary methods section 1). Response to first-line adjuvant chemotherapy was evaluated using available radiological data (supplementary methods section 1).

**Figure 1.**
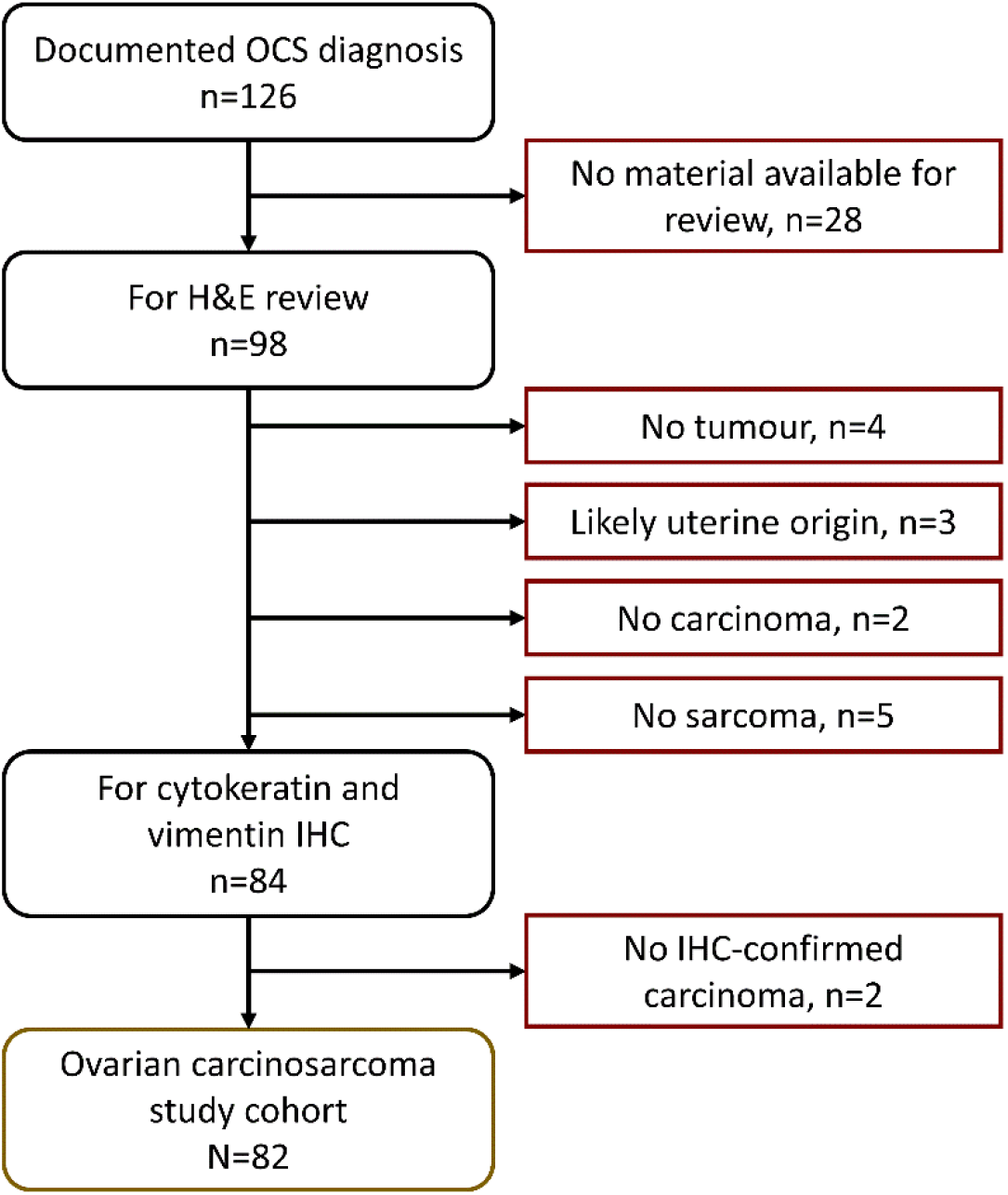
Case flow diagram for ovarian carcinosarcoma (OCS) cohort. IHC, immunohistochemistry. H&E, haematoxylin and eosin.

Ethical approval for the study was obtained from the Lothian NRS Human Annotated Bioresource (reference 15/ES/0094-SR1330) and the South East Scotland Cancer Information Research Governance Committee (reference CG/DF/E164-CIR21171). All participants gave written informed consent or had consent waived by the ethics committee due to the retrospective nature of the study.

### Pathology review

Of the 126 identified cases, archival formalin-fixed paraffin-embedded (FFPE) material was available for 98 cases. Pathology review was performed by an expert gynaecological pathologist (CSH) using H&E-stained slides from every available FFPE block. Available uterine samples were examined to confirm non-uterine origin (median 3 uterine blocks per case in the study cohort). A confirmatory observer (RLH) was present for all review.

Cases without a clear malignant high grade carcinomatous or sarcomatous component on H&E review were excluded (figure 1). Immunohistochemistry (IHC) for cytokeratins and vimentin were used to confirm the presence of both carcinomatous and sarcomatous compartments (1) (supplementary methods section 2); cases without IHC-confirmed carcinoma and sarcoma were excluded (figure 1).

Presence of endometriosis, squamous differentiation and serous tubal intraepithelial carcinoma (STIC) were documented as part of pathology review. The relative prevalence of carcinomatous and sarcomatous compartments across all available samples was documented (carcinoma-dominant: >70% carcinoma, sarcoma-dominant: >70% sarcoma, all others: mixed). Presence of carcinomatous and sarcomatous compartments in metastases (omentum and distant sites) was recorded during review (carcinoma only, sarcoma only, or mixed carcinosarcoma).

### Classification of carcinomatous and sarcomatous elements

The histotype of the carcinomatous compartment was determined by H&E review with IHC for WT1 and p53 in every case (supplementary methods section 2) (22). WT1 positivity was defined as positive tumour nuclei in carcinomatous cells. p53 staining was classified as aberrant-positive (aberrant diffuse nuclear positivity), aberrant-null (diffusely negative nuclei with confirmed adjacent positive stromal staining) or wild-type (variable nuclear positivity) (23).

Heterologous sarcomatous elements were identified from the H&E stained slides. Suspected chondrosarcoma and rhabdomyosarcoma were confirmed using IHC for S100 (chondrosarcoma: nuclear S100-positive) and desmin/myogenin (rhabdomyosarcoma: nuclear desmin/myogenin-positive) (supplementary methods section 2) (24). Liposarcoma was identified by the presence of adipocytes with malignant nuclei and was distinguished specifically from benign adipose tissue infiltrated by carcinosarcoma.

### HGSOC comparator cohort

A cohort of 362 otherwise unselected patients with a confirmed diagnosis of HGSOC following contemporary pathology review was used as a comparator cohort (supplementary methods section 3) (25). Characteristics of this cohort are summarized in supplementary table S1.

### Statistical analysis

All statistical analyses were performed using R version 4.1.0 (R Foundation for statistical Computing). Comparisons of frequency were made using the Chi-squared and Fisher’s exact test, as appropriate. Continuous data were compared using the Mann Whitney-U test. Survival analyses were performed using Cox proportional hazards regression models, presented as hazard ratios and respective 95% confidence intervals (HR, 95% CI). All tests were two-sided; P<0.05 was considered statistically significant.

### Statistical power

The statistical power of survival analyses were estimated using the powerSurvEpi R package. Power to detect a difference (HR=0.50) between two OCS populations within the study cohort (50:50 split) was 83.7% (using the study cohort survival event rate of 95.1%). Power to detect a survival difference (HR=0.50) against the HGSOC comparator cohort (N=362, event rate 90.1%) was 99.0%.

## RESULTS

### Patient characteristics

Of the 126 patients identified with a documented diagnosis of OCS, 82 were include in the study cohort (n=32 no tumour material available for pathology review, n=5 no sarcoma identified on H&E review, n=2 no carcinoma identified on H&E review, n=3 possible uterine origin, n=2 no confirmed cytokeratin-positive carcinoma) (figure 1). Baseline characteristics of the study cohort are summarised in table 1.

**Table 1.** Characteristics of ovarian carcinosarcoma cohort

|  |  | N | % / range |
| --- | --- | --- | --- |
| Cases | Total | 82 | - |
| Age | Median years | 69 | range 47-83 |
| FIGO stage at diagnosis | I | 9 | 11.4 |
|  | II | 8 | 10.1 |
|  | III | 51 | 64.6 |
|  | IV | 11 | 13.9 |
|  | NA | 3 | - |
| Neoadjuvant | Yes | 4 | 4.9 |
|  | No | 78 | 95.1 |
| First-line treatment | Platinum-taxane doublet | 26<sup>a</sup> | 31.7 |
|  | Other platinum combinations | 2 | 2.4 |
|  | Single-agent platinum | 26 | 31.7 |
|  | Other chemotherapy | 5 | 6.1 |
|  | Surgery only | 23<sup>b</sup> | 28.0 |
| Debulking status | No visible RD | 30 | 40.0 |
|  | Macroscopic RD<2cm | 15 | 19.0 |
|  | Macroscopic RD≥2cm | 30 | 38.0 |
|  | Macroscopic RD NOS | 4 | 5.1 |
|  | Unknown | 3 | - |
| Diagnosis period | Pre-1990s | 7 | 8.5 |
|  | 1990-1999 | 25 | 30.5 |
|  | 2000-2009 | 20 | 24.4 |
|  | 2010 onwards | 30 | 36.6 |
| Disease status | Relapsed | 75 | 91.5 |
|  | Stable at last follow-up | 7 | 8.5 |
| Vital status | Alive at last follow-up | 4 | 4.9 |
|  | Deceased - ovarian cancer | 72 | 87.8 |
|  | Deceased - other causes | 6 | 7.3 |

<sup>a</sup>1 with maintenance bevacizumab; <sup>b</sup>8 due to co-morbidities or performance status, 3 due to patient declining treatment, 3 due to early stage disease, 6 due to rapid deterioration following surgery, 3 unknown. FIGO, International Federation of Gynecology and Obstetrics; RD residual disease; NOS, not otherwise specified; FU, follow-up. NA, no available.

The majority of cases were FIGO stage III at diagnosis (51, 62.2% of evaluable cases; n=3 non-evaluable) (table 1). 9 (11.4%), 8 (10.1%) and 11 (13.9%) cases were FIGO stage I, II and IV. Median age at diagnosis was 69 years (range 47-83). 4 cases (4.9%) underwent neoadjuvant chemotherapy.

Median PFS was 9.6 months (95% CI 7.5-10.7). Median OS was 12.7 months (95% CI 9.2-17.1). For FIGO stage III/IV cases, the median OS was 11.9 months (95% CI 9.0-15.9).

### Histopathological characteristics of ovarian carcinosarcoma

The majority of OCS harboured an epithelial component of high grade serous (HGS) type (all confirmed WT1 positive) (n=65, 79.3%) (figure 2, figure 3); the remaining 17 cases (20.7%) were of endometrioid type (all confirmed WT1 negative). The vast majority of epithelial components had aberrant p53 expression patterns (wild-type pattern in 3 cases, 4.4% of evaluable carcinomatous components) (figure 2). The majority of cases did not demonstrate a dominance of either the sarcomatous of carcinomatous population (67.5%, 54 of 82 evaluable cases); 16 (20.0%) were carcinoma-dominant (>70% malignant cells of epithelial type); 10 (12.5%) were sarcoma-dominant (>70% malignant cells of mesenchymal type) (figure 2).

**Figure 2.**
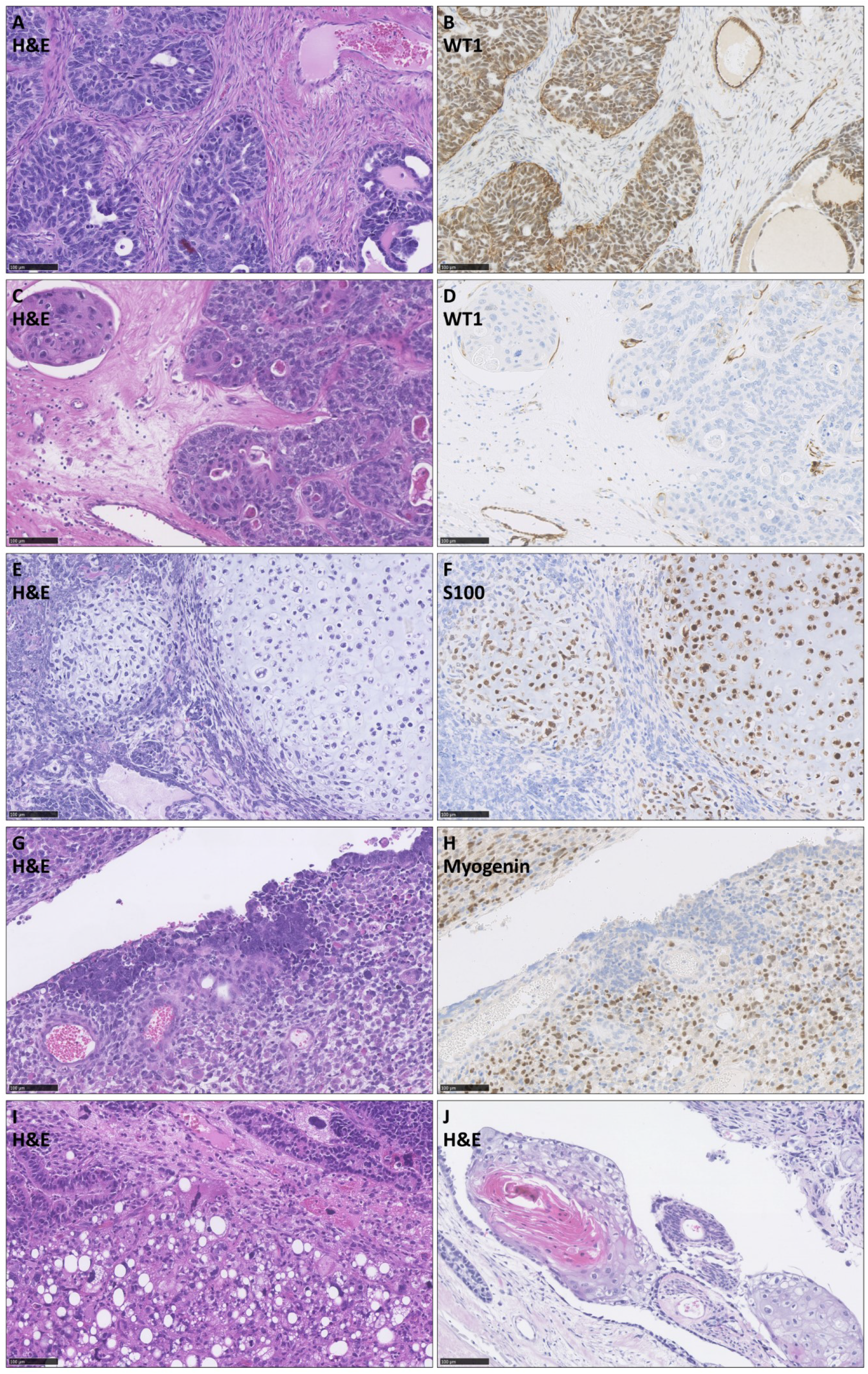
Histopathological features of ovarian carcinosarcoma (OCS). (A) and (B) WT1-positive epithelial component of high grade serous type. (C) and (D) WT1 negative epithelial component of endometrioid type. (E) and (F) OCS demonstrating S100-positive malignant cartilage. (G) and (H) OCS with myogenin-positive rhabdomyoblasts. (I) OCS demonstrating liposarcoma (confirmed cytokeratin-negative) adjacent to endometrioid epithelial component. (J) OCS demonstrating squamous differentiation. Scale bar represents 100µm.

**Figure 3.**
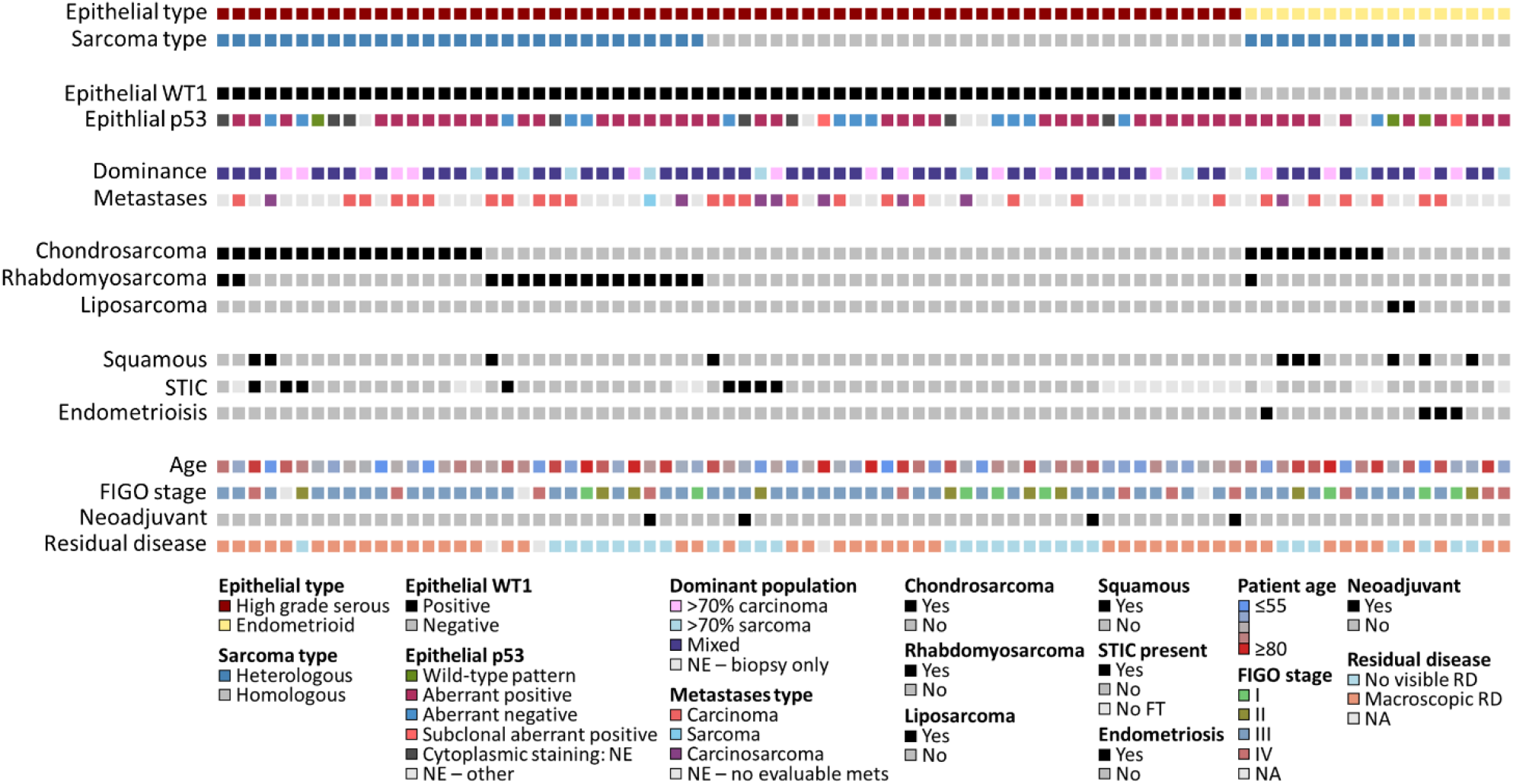
Clinical and histopathological landscape of ovarian carcinosarcoma. STIC, serous tubal intraepithelial carcinoma; FIGO, International Federation of Gynecology and Obstetrics; NE, non-evaluable; FT, fallopian tube; RD, residual disease; NA, not available.

Heterologous sarcomatous elements were identified in 42 cases (51.2%). The most common heterologous element was chondrosarcoma (31.7% of cases: 23 chondrosarcoma only, 3 chondrosarcoma plus rhabdomyosarcoma); rhabdomyosarcoma was also common (20.7% of cases; 14 rhabdomyosarcoma only, 3 chondrosarcoma plus rhabdomyosarcoma) (figure 2). Chondrosarcoma was significantly over-represented in OCS with endometrioid epithelial components (9 of 17, 52.9% in endometrioid vs 17 of 65 in HGS, 26.2%; P=0.044), with only one case demonstrating rhabdomyosarcoma (with concurrent chondrosarcoma) (figure 2). By contrast, rhabdomyosarcoma was common in OCS with HGS epithelial components (16 of 65 cases, 24.6%). Two OCS (2.4%) demonstrated liposarcoma: both had epithelial components of endometrioid type (P=0.041 for over-representation).

Endometriosis was identified in 4 cases, all of which had carcinomatous components of endometrioid type (figure 2). STIC lesions were identified in 8 cases, all of which had HGS epithelial components. Squamous differentiation was common in OCS with endometrioid carcinomatous components (35.3%, 6 of 17), but was also identified in 4 OCS with epithelial components of HGS type (6.2%, 4 of 65, including 1 case with an identified STIC).

The majority of cases with evaluable metastatic sites demonstrated metastases comprising only the carcinomatous population (75.0%, 27 of 36), with a minority showing mixed carcinosarcoma (22.2%, 8 of 36); metastasis of pure sarcomatous population was rare (2.8%, 1 of 36) (figure 2).

### Features associated with patient survival

OCS patients demonstrated similarly poor survival regardless of histological classification by carcinomatous (endometrioid vs HGS) or sarcomatous compartments (homologous vs heterologous) (figure 4A).

**Figure 4.**
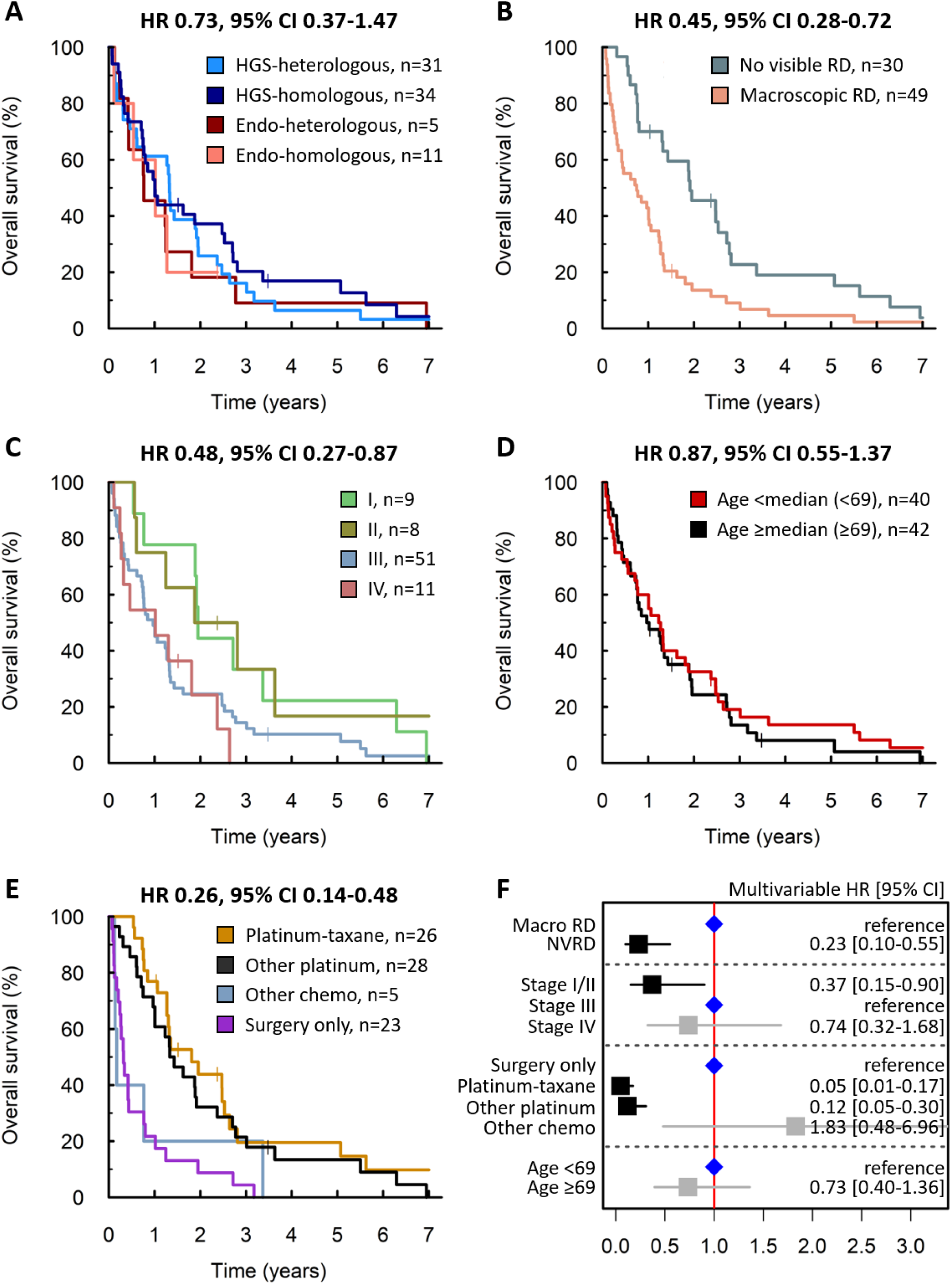
Ovarian carcinosarcoma patient survival. (A) Overall survival (OS) by histological subgrouping. Labelled hazard ratio (HR) reprints comparison of high grade serous (HGS) epithelial with homologous sarcoma (HGS-homologous) versus endometrioid (endo) epithelial with heterologous sarcoma (endo-heterologous); P=0.380. (B) OS by residual disease (RD) status following debulking surgery; P=0.001. (C) OS by stage at diagnosis. Labelled HR represents comparison of stage I/II versus stage III; P=0.015. (D) OS by age at diagnosis; P=0.538. Patients grouped according to median age at diagnosis across the cohort (69 years). (E) OS by first-line treatment regime. Labelled HR represents comparison of platinum-taxane versus surgery only; P<0.001. Additionally, HR for other platinum regimens (26 single-agent platinum, 2 other platinum combinations) versus surgery only is 0.34, 95% CI 0.19-0.61, P<0.001. (F) Forest plot of multivariable overall survival analysis, stratified by diagnosis period. Chemo, chemotherapy; macro, macroscopic; NVRD, no visible residual disease.

Achievement of no visible residual disease (NVRD) after surgical debulking was associated with significantly prolonged OS (HR=0.45, 95% CI 0.28-0.72) (figure 4B). Patients with stage I/II disease at diagnosis had significantly longer OS compared to stage III cases (HR=0.48, 95% CI 0.27-0.87) (figure 4C). Age at diagnosis did not have a significant impact on survival (figure 4D). Survival time was longest in patients who received platinum-taxane chemotherapy, but this was comparable to those receiving single agent platinum or other platinum combinations (figure 4E).

Multivariable analysis identified residual disease (RD) status, first-line treatment regime and stage as independently associated with survival (figure 4F).

### Comparison of ovarian carcinosarcoma with high grade serous ovarian carcinoma

OCS patients were significantly older at diagnosis compared to a comparator cohort of 362 unselected HGSOC patients (median 69 vs 61 years, P<0.0001) (figure 5A). A significantly greater proportion of OCS patients were diagnosed at FIGO stage I (2.7-fold enrichment; 11.4%, 9 of 79 evaluable OCS cases vs 4.3%, 15 of 351 evaluable HGSOC cases; P=0.025) (figure 5B). Differences in stage and age at diagnosis remained significant in a sensitivity analysis including only OCS with carcinomatous components of HGS type (P=0.033 and P<0.0001, respectively).

**Figure 5.**
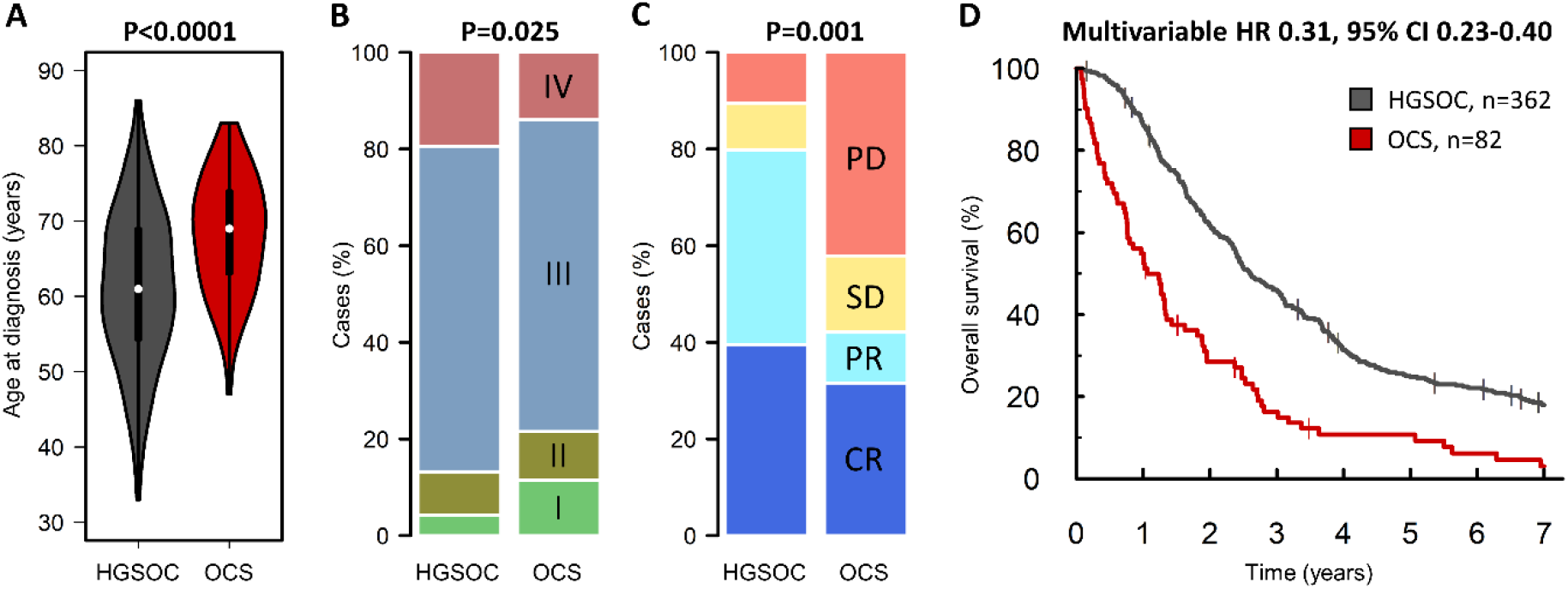
Comparison of ovarian carcinosarcoma (OCS) and high grade serous ovarian carcinoma (HGSOC). (A) Age at diagnosis. (B) FIGO stage at diagnosis; labelled hazard ratio P-value represents comparison of frequency of stage I cases. (C) Response to first-line chemotherapy; labelled P value represents comparison of overall response rate (complete response [CR] plus partial response [PR] versus stable disease [SD] plus progression disease [PD]). (D) Overall survival; labelled HR represents multivariable analysis of tumour type (HGSOC vs OCS; P<0.0001), age at diagnosis and stage at diagnosis, stratified by residual disease status.

Response rate to first-line adjuvant chemotherapy was significantly lower in OCS compared to HGSOC (42.1%, 8 of 19 evaluable OCS versus 79.8%, 99 of 124 evaluable HGSOC, P=0.001) (figure 5C). Multivariable analysis identified significantly shorter survival time for OCS patients compared to HGSOC (multivariable HR for HGSOC vs OCS 0.31, 95% CI 0.23-0.40, P<0.0001) (figure 5D). The difference in survival remained significant in a sensitivity analysis including only OCS who received platinum-containing chemotherapy (multivariable HR 0.42, P<0.0001).

## DISCUSSION

OCS are rare, biphasic malignancies that have received relatively little research attention to date. Although OCS is defined specifically as a biphasic tumour composed of high grade epithelial and mesenchymal components, the histopathological characteristics of OCS are highly heterogenous. Here we present a large OCS cohort with detailed clinical annotation and histopathological characterisation. To our knowledge, this is the largest pathologically-confirmed OCS cohort reported to date.

Most OCS cases demonstrated HGS epithelial components; however, OCS with endometrioid carcinomatous components also represented a major population (around 20% of cases). Smaller studies have shown the epithelial component is typically of HGS type, with other types representing only a minority of cases (1). In UCS, both serous-like and endometrioid-like UCS have been reported (26). Most OCS cases did not demonstrate a dominant malignant cell population, with only 20% and 12.5% of cases demonstrate >70% carcinomatous and >70% sarcomatous populations. The observation that the majority of metastases were of pure carcinomatous populations in line with data demonstrating that most metastases from UCS are carcinomatous (26).

Around half of OCS cases demonstrated heterologous sarcomatous elements. Overall, both chondrosarcoma and rhabdomyosarcoma were common, identified in 31.7% and 20.7% of cases, respectively. This is in contrast to UCS, where rhabdomyosarcoma is the most frequent heterologous element (approximately 20% of cases), with only around 10% of cases demonstrating chondrosarcoma (27). We observed a strong preference for chondrosarcoma over rhabdomyosarcoma in OCS with endometrioid epithelial components (rhabdomyosarcoma identified in only one case, co-occurring with chondrosarcoma). Liposarcoma was rare, consistent with its low frequency in UCS (<5%) (27), and was only observed in OCS demonstrating endometrioid epithelial components within our cohort. While squamous differentiation is typically an indicator of endometrioid carcinoma, and we show squamous differentiation in some OCS harbouring an endometrioid epithelial component, we also identified squamous differentiation in OCS cases with HGS carcinomatous compartments, indicating that squamous differentiation is not a specific feature of endometrioid tumours in this context. We identified endometriosis and STIC lesions in OCS with endometrioid and HGS carcinomatous components, respectively. This is consistent with the notion that OCS likely represent metaplastic carcinomas, and with endometriosis and STIC lesions being common precursor lesions of endometrioid and HGS ovarian carcinoma, respectively (28).

We demonstrate extremely poor survival in OCS patients, with high risk of relapse and death across patients of all stages, ages and first-line management strategies. The median overall survival time across the study cohort was 12.7 months. Earlier stage at diagnosis (FIGO I/II), undergoing platinum-containing adjuvant chemotherapy, and achievement of NVRD at debulking surgery were all associated with significantly prolonged survival upon multivariable analysis; however, mortality rate was still high in these patient groups. These findings are in line with the importance of optimal debulking across ovarian carcinoma types (21), and are in agreement with previous data suggesting improved survival in OCS with low residual disease volume (16-18). Previous reports of case numbers have suggested that presence of heterologous elements may be an indicator of poorer prognosis in OCS (29, 30), though other investigators have reported no significant association (31, 32). Histological subclassification of patients based on the carcinomatous and sarcomatous elements did not identify patient groups at higher or lower risk within our cohort, suggesting presence of heterologous elements is not of prognostic significance in OCS.

These data highlight the urgent need for improved treatment strategies for OCS patients. Some studies have investigated the potential role of ifosfamide in OCS management; ifosfamide-paclitaxel chemotherapy appears inferior to platinum-taxane regimens (33), while the relative efficacy of platinum-ifosfamide vs platinum-taxane remains controversial (1, 34, 35). Moreover, ifosfamide-containing may be less well tolerated (34, 36). The rarity of OCS has impeded progress of OCS-specific clinical trials, with OCS frequently included as a minor population alongside UCS. The GOG261 study of paclitaxel/ifosfamide versus carboplatin/paclitaxel demonstrated non-inferiority of the carboplatin-paclitaxel regime in a mixed cohort of UCS and OCS (36), though the majority of cases were UCS (>80%).

Progress toward discovery of effective molecularly targeted agents for OCS has been hindered by lack of molecular characterisation in this tumour type, and this has impeded inclusion of OCS in clinical trials based on specific molecular defects, agnostic to disease site (BASKET trials). Comprehensive molecular profiling to identify potentially actionable disease biology in OCS therefore represents an immediate research priority. Repurposing of drugs already in use for other cancer types represents a strategy that may facilitate rapid translation of candidate agents into early phase trials. International collaboration will be required to initiate disease-specific trials of OCS with sufficient power to inform future practice. Targeting of EGFR (37, 38), HER2 (37, 38), PDGFR (39, 40) and immunosuppressive molecules (41) have been suggested as potential strategies from molecular studies of gynaecological carcinosarcomas; however, data are limited and the vast majority of data are derived from UCS, rather than OCS. Molecular therapies routinely used for management of ovarian carcinoma may be of potential use in OCS; bevacizumab has demonstrated greatest efficacy in highest risk ovarian carcinoma cases (42), and may therefore be expected to benefit OCS patients, who are high risk by nature. Similarly, a minority of OCS are thought to harbour homologous recombination repair pathway defects (11), and may therefore be sensitive to PARP inhibition (43). Data regarding clinical efficacy of anti-angiogenic agents and PARP inhibitors in OCS are extremely limited. A phase II trial of the anti-angiogenic agent aflibercept demonstrated disappointing activity in recurrent gynaecological carcinosarcoma (44); however, this study included only three OCS cases.

While OCS were originally considered separate to epithelial ovarian cancer, it is now believed that OCS represent metaplastic carcinomas (3). This has led many to consider OCS as variants of HGSOC (45): both are commonly diagnosed at advanced stage, together representing the most aggressive ovarian cancer types, and the majority of OCS harbour carcinomatous components of HGS type. However, we demonstrate that OCS are around three times more likely to be diagnosed at FIGO stage I, are significantly older at diagnosis (median 69 years), show significantly greater levels of intrinsic chemoresistance (response around 40%) and demonstrate significantly shorter survival compared to an unselected HGSOC population (multivariable HR for HGSOC 0.31). Moreover, a significant proportion of OCS have an epithelial component of endometrioid type. These data suggest that consideration of OCS as variants of HGSOC is a substantial over-simplification, and that OCS in fact represent a distinct high-risk ovarian cancer type with unique clinical behaviour and histopathological characteristics. For forthcoming trials where OCS may be included alongside high grade endometrioid and HGS ovarian carcinoma, appropriate stratification is recommended.

Major strengths of this work include contemporary pathology review of a large cohort of cases (n=82), exclusion of cases with uterine origin, and the use of IHC to confirm presence of both carcinomatous and sarcomatous populations, to confirm the histotype of the carcinomatous elements, and to confirm the presence of chondrosarcoma and rhabdomyosarcoma. The majority of OCS studies have reported only a small number of cases (typically fewer than 30) (1) with limited pathological assessment. The detailed clinical annotation and mature outcome data (event rate >90%) available for our cases - prospectively collected as part of routine care - is another major strength, alongside the use of a pathologically-confirmed HGSOC comparator cohort. Limitations include the retrospective nature of the study, and the extensive study period, with guidelines for ovarian cancer management evolving over this time. However, a long study period was essential for curating a sufficient number of cases for meaningful analysis, which has represented a significant obstacle in previous studies.

## CONCLUSION

OCS represents an extremely aggressive form of ovarian cancer, with distinct clinical behaviour compared to HGSOC. OCS patients are poorly served by currently available treatment options, and new therapeutics strategies – which have been hindered by lack of research attention and the relative rarity of OCS – are urgently required to improve patient outcomes. Absence of RD following debulking surgery and earlier stage at diagnosis are markers of improved survival; however, risk of recurrence and mortality is high across all patient populations. While OCS are histopathologically heterogeneous, significant relationships exist between phenotypes of the carcinomatous and sarcomatous compartments.

## Data Availability

We are happy to provide relevant data upon reasonable request, subject to compliance with the relevant ethical framework.

## Acknowledgements

We thank the patients who contributed to this study and the Edinburgh Ovarian Cancer Database from which the clinical data reported here were retrieved.

## Author contributions

RLH: conceptualisation, formal analysis, investigation, methodology, visualisation, funding acquisition, writing – original draft. IC: investigation, writing – review and editing. MC: data curation, project administration, writing – review and editing. TR: data curation, investigation, writing – review and editing. CB: data curation, writing – review and editing. CG: supervision, resources, writing – review and editing CSH: investigation, methodology, supervision, writing – review and editing.

## Ethics approval and consent to participate

Ethical approval for the study was obtained from the Lothian NRS Human Annotated Bioresource (reference 15/ES/0094-SR1330). All participants gave written informed consent or had consent waived by the ethics committee due to the retrospective nature of the study. The study was performed in accordance with the Declaration of Helsinki.

## Consent for publication

Not applicable.

