## supplementary materials for "Ovarian carcinosarcoma is a distinct form of ovarian cancer with poorer survival compared to tubo-ovarian high grade serous carcinoma"

### SUPPLEMENTARY INFORMATION

#### Supplementary methods

*1. Evaluation of therapy response and disease progression*

Response to first-line chemotherapy was evaluated using measured changes in disease volume by radiological assessment. 63 cases were non-evaluable for first-line response (23 no adjuvant chemotherapy, 14 no disease to follow on scan, 19 insufficient radiological investigations, 7 measurements not reported on scan). Complete response was defined as complete resolution of pre-treatment disease. Partial response (PR) was defined as disease reduction by ≥50%. Progressive disease (PD) was defined as appearance of new radiologically-confirmed lesions or ≥50% increase in measured disease volume. Evaluable cases not reaching criteria for PR or PD were classified as stable disease (SD).

Progression-free survival (PFS) was calculated as time from pathologically confirmed diagnosis to recurrence/progression as determined by radiology (n=31, 37.8%), CA125 tumour marker (n=10, 12.2%), positive biopsy (n=2, 2.4%), or physician-determined progression in the absence of other investigations (n=23, 28.0%); 9 cases (11.0%) were non-evaluable for progression.

*2. Immunohistochemistry*

Pan-cytokeratin and vimentin immunohistochemistry (IHC) was performed with Leica BOND ready-to-use AE1/3 and vimentin antibodies on the Leica BOND III Autostainer with IHC protocol F. High grade serous ovarian carcinoma (HGSOC) (cytokeratin positive, vimentin negative) and normal ovarian stroma (cytokeratin negative, vimentin positive) were used as controls.

WT1 IHC used 1:1000 monoclonal 6F-H2 antibody (Dako); p53 IHC used 1:50 monoclonal DO7 antibody (Sigma). HGSOC was used as a positive control for WT1 (1). HGSOC with missense *TP53* mutation and

HGSOC with frameshift *TP53* mutation were used as controls for the aberrant-positive and aberrant-null p53 patterns, respectively. Non-malignant stroma was used as a control for wild-type staining.

IHC for estrogen receptor (ER) (1:50 monoclonal antibody M3643 clone EP1) and Napsin A (Leica ready-to-use antibody) was used to distinguish endometrioid carcinoma from potential clear cell carcinoma in one case. Low grade serous ovarian carcinoma and clear cell ovarian carcinoma were used as positive controls for ER and Napsin A, respectively.

S100, desmin and myogenin (Myf4) IHC was performed with Leica BOND ready-to-use antibodies on the Leica BOND III Autostainer using IHC protocol F. HGSOC and non-malignant stroma were used as negative controls. Known S100 chondrosarcoma (via routine NHS pathology) was used as a positive control for S100. Known desmin-positive rhabdomyosarcoma (via routine NHS pathology) was used as a positive control for desmin and myogenin.

*3. High grade serous ovarian carcinoma comparator cohort*

A cohort of 362 pathologically-confirmed HGSOC cases were identified as part of a previous study, and used as a comparator cohort (supplementary table 1). Briefly, cases were identified following pathology review of tumour material from 539 unselected patients with a diagnosis of ovarian, peritoneal or fallopian tube carcinoma treated at the Edinburgh Cancer Centre. All patients received platinum-containing adjuvant chemotherapy after primary debulking surgery. Outcome data and therapy response data were reported to the same criteria as for the OCS cohort. For a full description of HGSOC cases, see reference (2) below.

**Supplementary tables**

Supplementary table S1. Characteristics of high grade serous ovarian carcinoma comparator cohort (N=362).

| | | N | % |
|---|---|---|---|
| **Age at diagnosis** | Median years | 61 | Range 33-86 |
| **FIGO stage** | I | 15 | 4.3 |
| | II | 31 | 8.8 |
| | III | 237 | 67.5 |
| | IV | 68 | 19.4 |
| | NA | 11 | - |
| **Vital status at last follow-up** | Alive | 36 | 9.9 |
| | Deceased | 326 | 90.1 |
| **Median OS** | Years | 2.60 (95% CI 2.40-3.07) | |

FIGO, International Federation of Gynecology and Obstetrics; NA, not available
